# Privacy-Protecting, Reliable Response Data Discovery Using COVID-19 Patient Observations

**DOI:** 10.1101/2020.09.21.20196220

**Authors:** Jihoon Kim, Larissa Neumann, Paulina Paul, Michael Aratow, Douglas S. Bell, Jason N. Doctor, Ludwig C. Hinske, Xiaoqian Jiang, Katherine K. Kim, Michael E. Matheny, Daniella Meeker, Mark J. Pletcher, Lisa M. Schilling, Spencer SooHoo, Hua Xu, Kai Zheng, Lucila Ohno-Machado, for the R2D2 Consortium

**Author notes:** R2D2 Consortium authors are listed in the Supplementary Materials. These authors contributed equally to this work (Jihoon Kim, Larissa Neumann).

## Abstract

There is an urgent need to answer questions related to COVID-19’s clinical course and associations with underlying conditions and health outcomes. Multi-center data are necessary to generate reliable answers, but centralizing data in a single repository is not always possible. Using a privacy-protecting strategy, we launched a public *Questions & Answers* web portal (https://covid19questions.org) with analyses of comorbidities, medications and laboratory tests using data from 202 hospitals (59,074 COVID-19 patients) in the USA and Germany. We find, for example, that 8.6% of hospitalizations in which the patient was not admitted to the ICU resulted in the patient returning to the hospital within seven days from discharge and that, when adjusted for age, mortality for hospitalized patients was not significantly different by gender or ethnicity.

**One Sentence Summary:** Publicly Sharing Knowledge on COVID19 Without Sharing Patient-Level Data: A Privacy-Protecting Multivariate Analysis Approach

## Main Text

The severe acute respiratory syndrome coronavirus 2 (SARS-CoV-2) pandemic represents a watershed event in public health and has highlighted numerous opportunities and needs in clinical and public health informatics infrastructure (*1*–*3*). One of the key challenges has been the rapid response of analyses and interpretation of observational data to inform clinical decision making and patient expectations, understanding, and perceptions (*4*–*8*).

Several initiatives are building COVID-19 registries or consortia to analyze electronic health record (EHR) data (*6, 7, 9*). The expectation is that these resources will provide researchers and clinicians access to a rich source of observational data to understand the clinical progression of COVID-19, to estimate the impact of therapies, and to make predictions regarding outcomes.

Registries may contain limited data for patients diagnosed with COVID-19: the barriers for having more data are based on both privacy concerns and also on what elements have been deemed valuable by health professionals and researchers at a particular point in time. The problems with a new and evolving disease like COVID-19 is that we do not know what data or information will be most valuable. For example, in the pandemic’s early stages, the dermatological and hematological findings were not evident, and those data were not included in registries or reports. Interest in specific laboratory markers (e.g., D-dimer, troponin) for these disturbances and additional symptoms (e.g. anosmia, conjunctivitis) has increased over time.

Additionally, it is challenging for researchers and clinicians to understand the structure and quality of the data in registries and other types of data repositories, and to formulate queries to consult the data in their institution. The process becomes more complicated when data from multiple institutions are involved.

Thus, the utilization of EHRs to characterize COVID-19 disease progression and outcomes is challenging. However, observational studies using EHR data may be useful when a research question does not lend itself to a randomized clinical trial (RCT). Observational studies may also help determine if results from RCTs replicate after relaxing eligibility criteria for real-world applications. While the scientific community has raised concerns about the reproducibility of findings, data provenance, and proper utilization of observational data, resulting in some COVID-19 articles being retracted (*10*), there remains a clear need to responsibly, ethically, and transparently analyze observational data to provide hypothesis generation and guidance in the pursuit of evidence-based healthcare.

We focus on using novel decentralized data governance and methods to analyze EHR-derived data. Researchers’ questions posed in natural language are adapted to standardized queries using distributed data maintained in 12 health systems, covering 202 hospitals located in all U.S. states and two territories, and one international academic medical center (Table 1). This collaboration provides the capability to answer questions that require comparisons with historical data from over 45 million patients and uses a dynamic approach to account for an evolving awareness of the most impactful COVID-19 questions to answer and hypotheses to explore. Having access to complete EHR data from 10 of these health systems (two sites use COVID-19 registries), and not just a predefined list of key data elements, differentiates our approach from centralized registries and public health reports. The ability to build and evaluate multivariate models across a large number of health systems and to integrate results from registries differentiates our approach from most federated clinical data research network approaches.

**Table 1.** Participating sites. Cedars Sinai Medical Center (CSMC), University of Colorado Anschutz Medical Campus (CU-AMC), Ludwig Maximillian University of Munich (LMU), San Mateo Medical Center (SMMC), University of California (UC) Davis (UCD), Irvine (UCI), San Diego (UCSD), San Francisco (UCSF), University of Southern California (USC), University of Texas Health Science Center at Houston and Memorial Hermann Health System (UTH), Veterans Affairs Medical Center (VAMC). *Available data on hospital characteristics from 2018.

| Institution | Hospitals | Beds | Discharges<br>per year | EHR system | Data Source |
| --- | --- | --- | --- | --- | --- |
| CSMC | 2 | 1,019 | 61,386 | Epic | EHR |
| CU-AMC | 12 | 1,829 | 106,325 | Epic | EHR |
| LMU* | 12 | 1,964 | 78,673 | SAP/i.s.h.med<br>QCare<br>IMESO | COVID-19<br>Registry |
| SMMC | 1 | 62 | 1,951 | Harris Software<br>(Pulsecheck)<br>Cerner (Soarian)<br>eClinicalworks | EHR |
| UCD | 1 | 620 | 32,248 | Epic | EHR |
| UCI | 1 | 417 | 21,656 | Epic | EHR |
| UCLA | 2 | 786 | 47,491 | Epic | EHR |
| UCSD | 3 | 808 | 29,895 | Epic | EHR |
| UCSF | 3 | 796 | 48,120 | Epic | EHR |
| USC | 2 | 1,511 | 23,454 | Cerner | EHR |
| UTH | 17 | 4,164 | 233,890 | Cerner | COVID-19<br>Registry |
| VAMC | 146 | 13,000 | 676,402 | ViSta/CPRS | EHR |
| Total | 202 | 26,976 | 1,361,491 |  |  |

The responsibility of translating the question into code and of performing quality control processes lies among members of the Reliable Response Data Discovery for COVID-19 Clinical Consults using Patient Observations (R2D2) Consortium (see supplemental materials). The analyses do not require data transfer outside these institutions and reduce the risk of individual or institutional privacy breaches. We produce results that are publicly available as soon as they pass quality controls. In this approach, there is targeted analysis of specific data elements at a local level. Only the results of calculations (e.g., counts, statistics, coefficients, variance-covariance matrices) performed on data transformed into the Observational Medical Outcomes Partnership Common Data Model (OMOP CDM) from relevant patient cohorts are released from the healthcare institutions; no individual patient-level data are shared (*11*).

Between December 11, 2020 and August 31, 2020, our consortium had 928,255 tested patients for SARS-CoV-2, 59,074 diagnosed with COVID-19, with 19,022 hospitalized and 2,591 deceased. Our public Questions and Answers (Q&A) portal (https://covid19questions.org) provides answers to research questions using several univariate or multivariate analyses, including potential associations between mortality and comorbidities; pre-hospitalization use of anti-hypertensive medications; laboratory values and hospital events. For each question, we report on the number of participating institutions and the time period within which local queries were run. Figure 1 shows the graphical display of two answers.

**Fig 1.**
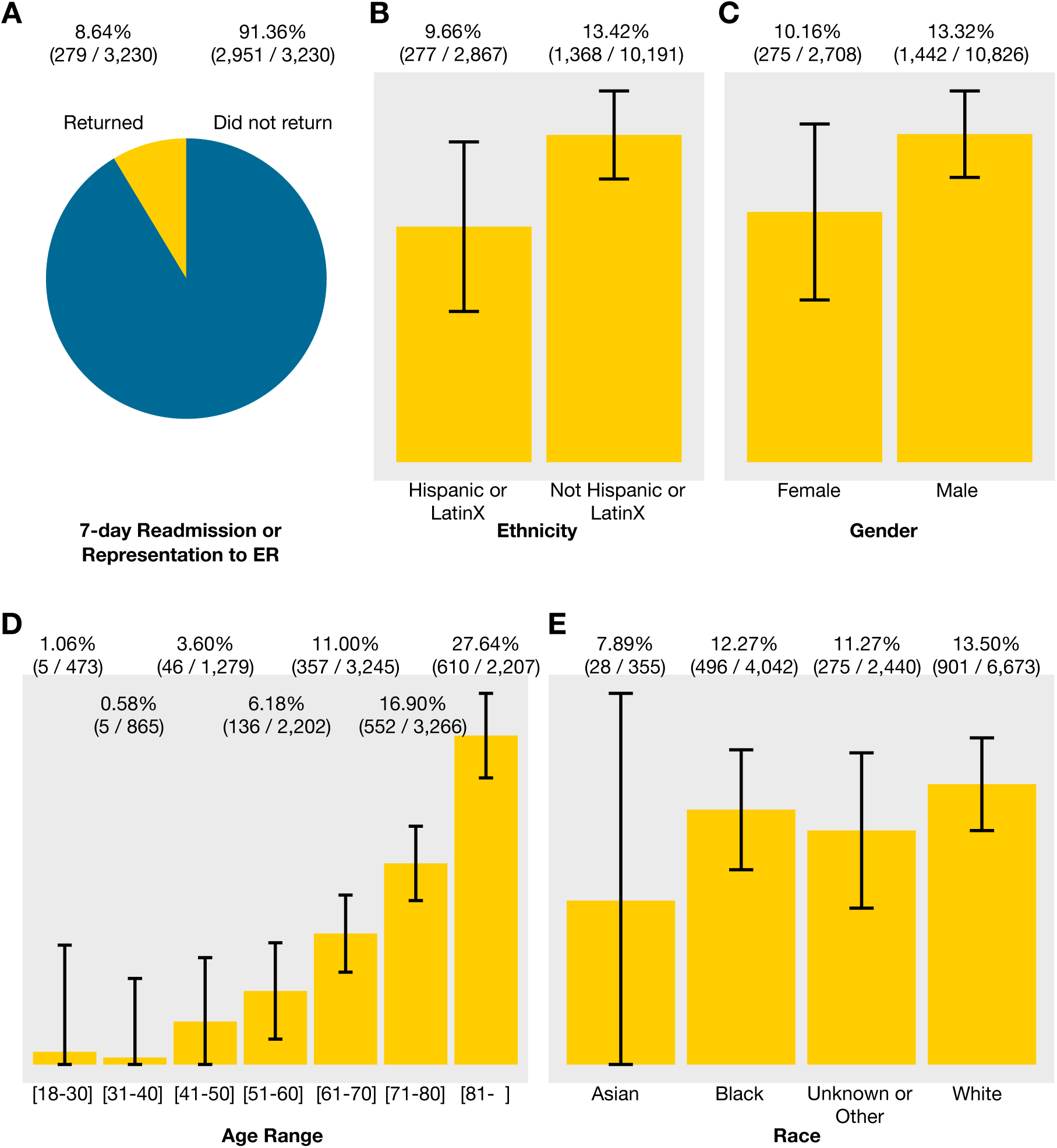
Examples of two COVID-19 *Questions and Answers*: Return to hospital and mortality. (A) 8.6% of hospitalizations without an ICU admission resulted in the patient presenting to the Emergency Room or a hospital readmission within seven days (data from ten health systems). (B-E) Unadjusted mortality rates from aggregated results are shown with 95% confidence intervals (data from ten health systems). Univariate analyses indicate that lower *age, Hispanic ethnicity*, and *female gender* are associated with lower mortality for adult hospitalized COVID-19 patients.

**Example 1**. “*Many adult COVID-19 patients who were hospitalized did not get admitted to the ICU and were discharged alive. How many returned to the hospital within a week, either to the Emergency Room (ER) or for another hospital stay?*” The answer indicates 8.6%. This question is both important from the standpoint of understanding the natural course of disease and planning for needed resources. Although efforts are underway to understand post-discharge outcomes in COVID-19 infected patients, to date they have been limited to case series (*12*), modest sample sizes (*13*), or single-center or geographically concentrated health systems (*14, 15*). These extant studies may also be hampered by fixed inclusion/exclusion criteria (*16*).

**Example 2**. “*Among adults hospitalized with COVID-19, how does the in-hospital mortality rate compare per subgroup (age, ethnicity, gender and race)?*” The answers from univariate and multivariate analyses (logistic regression, Fig. 2) indicate that *age* is a major risk factor, but *ethnicity* and *gender* are also significant when considered univariately. There is great interest and growing peer-reviewed literature on risk factors for COVID-19 mortality: the agility of our approach allows us to quickly re-run queries and rebuild models as new predictors become relevant and the understanding of the disease evolves (*17*–*20*).

**Fig 2.**
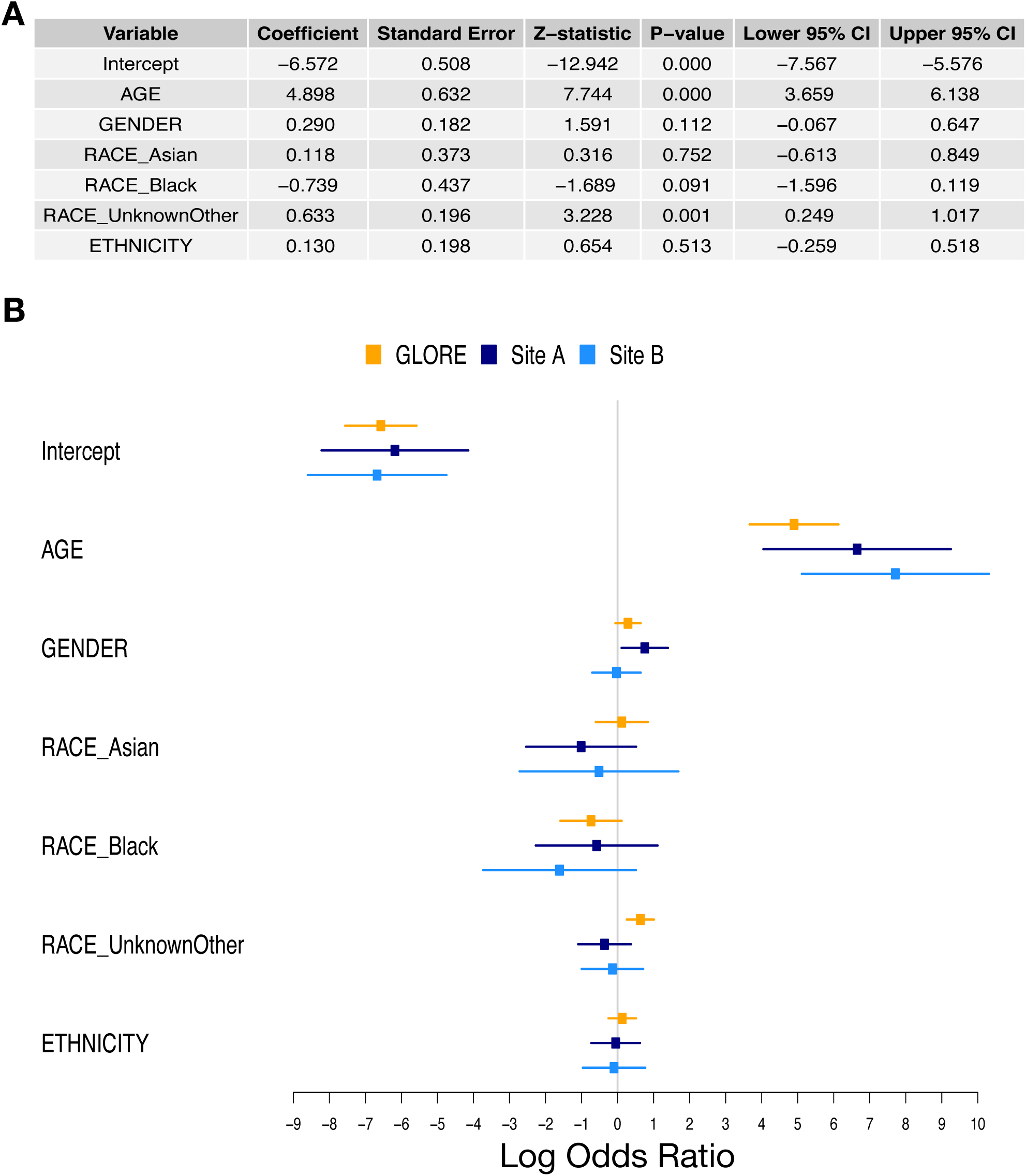
Regression Results. (A) Adjusted effects from the Grid Binary LOgistic REgression (GLORE) *(11)* federated logistic regression model (3,146 patients from eight health systems). The baselines were GENDER=*female*, RACE=*white*, ETHNICITY=*non-hispanic. Age* (in years) was divided by 100. After adjustment via distributed logistic regression, *age* remains significant. (B) Results from local logistic regression performed at two sites are also shown for comparison with GLORE results.

Several other questions and answers are shown in the portal and are updated when answers are approved for posting. We also have a set of approved questions awaiting response from the majority of sites.

Institutions participating in our network are diverse in terms of organization, population served, prior investments in information technology, and location. A novel governance structure in our consortium allows us to distribute the workload across various teams without relying on a traditional coordinating center, instead including a Consortium Hub for certain functions.

This approach keeps patient data in-house, simplifies data use agreements, avoids delegation of control of patient data to another institution, and allows any institution to benchmark its results to those produced by the consortium, since all questions and respective final, aggregated answers, database query code, concept definitions and analytics code are made public. It complies with HIPAA, the Common Rule, the GDPR, and the California Consumer Privacy Act with regards to handling of patient data. Code sharing and public answers promote transparency and reproducibility without disclosing patient information or institutional information.

Our approach has advantages but also some limitations. The advantages are that we are able to, in relatively short time, publicly post answers to questions that are of general interest, using data from a spectrum of highly diverse institutions with different levels of information technology baselines and expertise in standardized data models and vocabularies, institutional policies, state and federal regulations. Because we keep data locally and only consult data elements that are necessary to answer specific questions, this approach has a lower risk of privacy breach when compared to registries in which patient data are exchanged or to distributed consortia in which summary-level results for each institution are reported. Additionally, since registries typically focus on a single disease or condition, they often lack comparator data from other patients, limiting the opportunity to characterize a new disease and discover how it differs from what we currently know. Participating sites do not need to transform all data into OMOP CDM and can decline to answer any questions they do not feel comfortable with or answer partially to ensure patient-level privacy by masking counts between 1 and 10. Institutional privacy is also preserved because all public answers combine the aggregate data from at least three Responding Sites (we do not specify which ones), but we keep all answers for audits. Making concept definitions, query code, and results available allows reproducibility and enables automated updates to the answers. A major advantage is that existing registries of consortia can serve as additional sites to help answer certain questions.

The limitations are inherent from considering all sites equal when formulating a final answer. Regional or institutional practice variations are not represented in the answers. Additionally, the distributed nature of the R2D2 consortium adds a requirement for a dynamic management team, the need to educate local leadership on distributed analytics, and potential for delays in certain decisions in order to exercise shared governance. A specific limitation of our current consortium is the preponderance of institutions based in California: eight out of 12 (67%), accounting for 17.5% of COVID-19 patients (Fig. 3). This was a convenience sample of organizations that had shared interests and a history of collaboration. We invite other institutions, consortia and registries worldwide to join us in answering questions of general interest.

**Fig 3.**
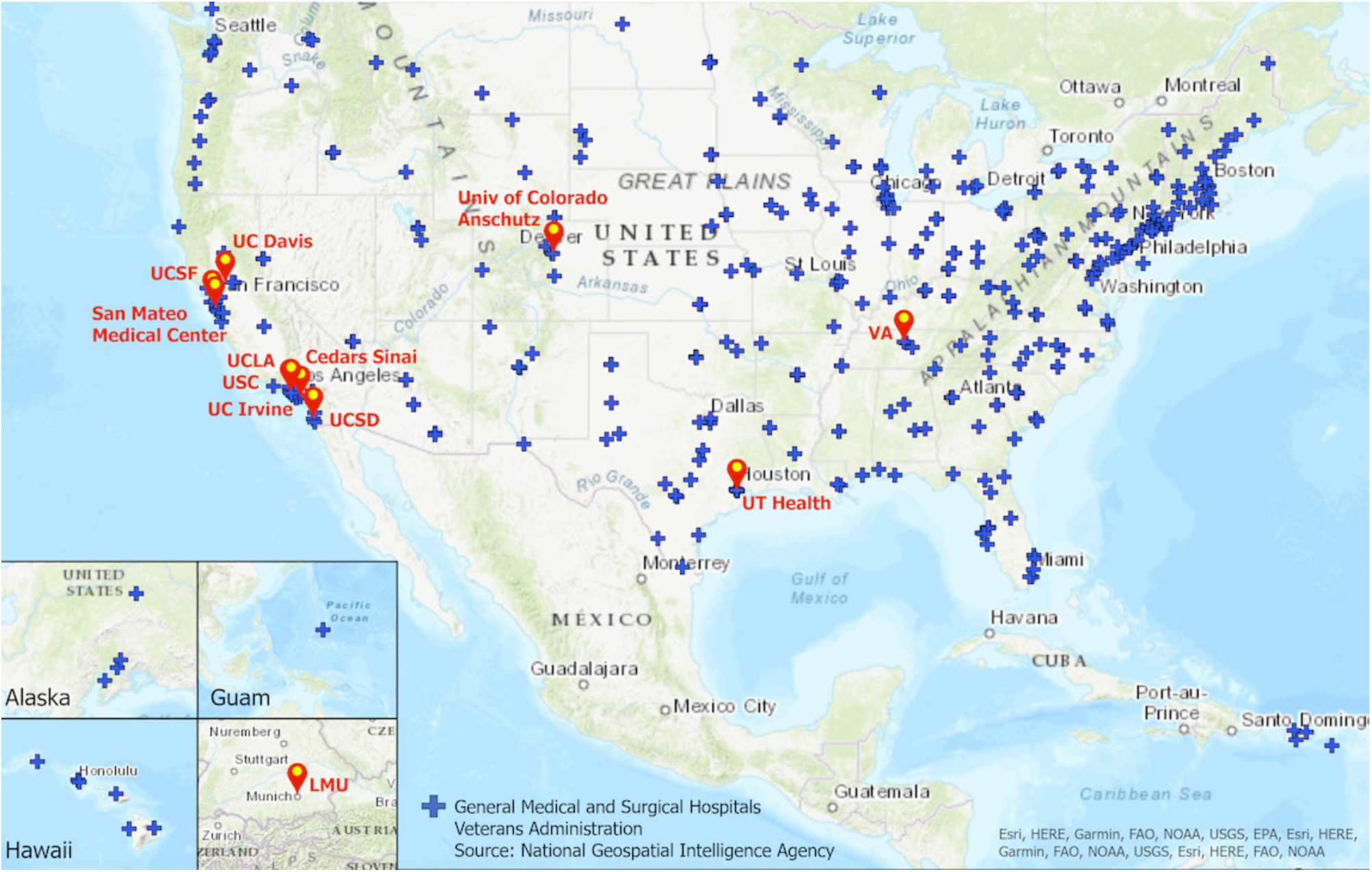
Location of consortium’s medical centers and hospitals. Map by Ilya Zaslavsky

## Data Availability

Answers are available at https://covid19questions.org and code is in https://github.com/DBMI/R2D2-Public.

## Acknowledgments

We thank Dr. Michael Hogarth for insightful comments.

## Author contributions

JK, LN and LO-M drafted the manuscript. PP, MA, DB, JD, LH, KK, DM, MM, MP, LS, SS, HX, KZ organized local teams for answering questions. All authors reviewed the manuscript. LO-M supervised the study and acquired the funding. See supplemental materials for a complete list of consortium authors;

## Supplementary Materials

Materials and Methods

Figs. S1 to S5

Tables S1 to S5

R2D2 Consortium Members

